# Emergence of Severe Metastatic Hypervirulent *Klebsiella pneumoniae* infections in Texas

**DOI:** 10.1101/2021.06.08.21257347

**Authors:** Junaid M. Alam, Haris Ahmed, Hong M. Thai, Kevin Garnepudi, Ramesh B. Kesavan, Gnananandh Jayaraman, Anna Sangster, Dylan Curry, Heidi A. Butz, Lori Smith, Maureen Vowles, Kelly F. Oakeson, Erin L. Young, Siva T. Sarva

## Abstract

**Background:** Hypervirulent Klebsiella pneumoniae (hvKp) infections have mainly been described in Asia. Two patients in the age group of 30 to 50 years presented within a two month period to a tertiary referral hospital in Texas with septic shock, hepatic abscess and septic thrombophlebitis. Blood cultures were positive for *Klebsiella pneumoniae* (isolates 2020CK-00441 and 2021CK-00720 respectively).

**Methods:** Whole genome sequencing was performed using paired-end Illumina MiSeq reads for both isolates. Nanopore sequencing to obtain a closed genome was performed for 2020CK-00441.

**Results:** 2020CK-00441 belonged to ST23 type while 2021CK-00720 was a ST65 type isolate. Kleborate analyses predicted with high confidence that both the isolates were hvKp. Phylogenetic analyses showed that the two strains are not closely related to each other or to any other known hvKp isolates. Both the isolates had yersiniabactin, colibactin, aerobactin and salmochelin producing loci which likely confer these isolates hvKp phenotype. 2020CK-00441 had a unique pK2044 like plasmid.

**Conclusions:** HvKp strains capable of causing devastating metastatic septic infections have emerged in Texas. These isolates are unique when compared to other hvKp strains of the world. Country wide surveillance and whole genome sequencing of these strains is essential to prevent a major public health emergency in USA.

## INTRODUCTION

Hypervirulent *Klebsiella pneumoniae* (hvKp) was first recognized in Taiwan in 1986 [1]. HvKp unlike the classical *Klebsiella pneumoniae* (cKp) affects healthy immunocompetent patients in community settings rather than being acquired as a healthcare associated infection [2, 3]. HvKp infection presents with fever, abdominal pain, and multiple hepatic abscesses and often metastasizes to multiple body sites, leading to septic emboli, endophthalmitis, brain abscess, and epidural abscess. While most cases of hvKp are reported in Southeast Asia, it is an emerging pathogen in the Western countries [4–8]. Multi drug resistant and extensively drug resistant hvKp strains are emerging around the world [9–12].

Initially it was believed that hypermucoviscous phenotype identified by a “string test” was diagnostic of hvKp [13]. However, it was recently shown that many hvKp isolates are not hypermucoviscous and also that a few cKp isolates are hypermucoviscous suggesting that the test is unreliable for characterizing hvKp [14, 15]. Furthermore, due to the presence of K1 and K2 capsule types in the hvKp isolates, capsule type alone can not be used to predict hvKp [16].

Klebsiella pneumoniae has been classified into many sequence types (ST types) using, multi-locus sequence typing (MLST) and various clonal groups (CG groups) based on whole genome (WGS) sequencing [3]. Putative virulence factors, a) plasmid associated loci, iro (salmochelin biosynthesis), iuc (aerobactin synthesis), rmpA (regulator of mucoid phenotype) b) Integrative and Conjugative element *Klebsiella pneumonia* (ICEKp) associated loci yersiniabactin and colibactin, are found to be more prevalent in hvKp isolates [3, 15].

HvKp has the potential to become a major public health problem. To this end isolates of the *Enterobacterales* order, carrying one or more genetic markers of hypervirulence including: peg-344, iroB, iucA, rmpA, and rmpA2 have recently been added to the list of AR/HAI alerts for public health laboratories within the Antibiotic Resistance Laboratory Network (ARLN). There is no consensus regarding the clinical and genetic characteristics that determine an isolate to be hvKp. Large scale studies in the United States thus far have only focused on limited genomic analyses of the available isolate collections [17, 18]. Whole genome sequences of clinical isolates of hvKp in USA have been described before but they do not appear to be isolated from patients with life threatening metastatic infections [19, 20]. A clear enumeration of the clinical presentation, whole genome sequencing and epidemiological analyses of the prevalent hvKp isolates in United States is needed. To the best of our knowledge, we report for the first time in USA, two whole genome sequences of hvKp which caused severe metastatic life-threatening septic infections.

## METHODS

### Clinical Cases

Consent was obtained from the patient and/or patient family to publish the clinical findings of the patients. The study was deemed to be IRB exempt and need for IRB oversight waived by C.A.R.R.I.E. (Centralized Algorithms for Research Rules on IRB Exemption) of HCA Health Care and also by local research review committee of HCA Houston Health Care, Kingwood.

### Isolate preparation and antibiotic sensitivity determination

Blood cultures were obtained using standard blood culture techniques in conjunction with Bactec FX system. Antibiotics sensitivities were obtained using both the Microscan Walkaway system and Sensititre GNX2F panels (Thermo-Fisher Scientific). Additionally, the modified carbapenem inactivation method (mCIM) was used as a phenotypic method to assess for carbapenemase production. Details of these methodologies are provided in the supplemental data.

### Whole genome sequencing and bioinformatics

Bacterial isolates were grown on trypic soy agar with 5% sheep blood overnight. DNA was extracted using the Qiagen EZ1 DNA Tissue Kit (953034). WGS for 2020CK-00441 was done on both the Illumina MiSeq with Nextera DNA flex library preparation kits (20018705) [21], and Oxford Nanopore GridION with the Ligation Sequencing kit for the flongle adapter (FLO FLG001;SQK-LSK109) in accordance to manufacturer’s instructions. Fastq files are available from the SRA database as SRR13075498 and SRR13436874 respectively. A closed genome for 2020CK-00441 was obtained by utilizing a hybrid assembly of both long reads and short reads using Unicycler [22], and annotated with PGAP.

2021CK-00720 only underwent short-read Illumina sequencing. Fastq files are available from the SRA database as SRR13965555. Fastq reads were filtered for quality with Seqyclean a comprehensive preprocessing software pipeline [23]. Cleaned reads were *de novo* assembled with shovill (unpublished)(https://github.com/tseemann/shovill) into contigs. Bioinformatic hypervirulence prediction was performed via kleborate, a tool to screen genome assemblies of *Klebsiella pneumoniae complex*.

(https://www.biorxiv.org/content/10.1101/2020.12.14.422303v2).

### Genome and Plasmid Comparisons

2020CK-00441 and 2021CK-00720 were compared to each other and the hypervirulent Klebsiella genomes downloaded from BioProjects PRJNA506506, PRJNA509089, PRJNA509091, PRJEB38367, PRJNA349219, PRJNA391211, PRJNA638288 and PRJEB34922. ST23 and ST65 typing was ascertained from kleborate, and contig and genome fasta files were annotated with Prokka and aligned with Roary, a pan genome pipeline, into a core genome alignment [24, 25]. Snp-dists software was used to count the number of SNPs between the two described isolates (unpublished)(https://github.com/tseemann/snp-dists). Phylogenetic trees were generated with iqtree and visualized with ggtree. [24–27] The top 10 blast hits for p2020CK-00441_1were annotated with prokka plus NC_006625/NTUH-K2044 plasmid pK2044 and aligned with clinker to visualize synteny[28].

## RESULTS

### A. Clinical Presentation

#### a) Case #1

A patient in the age group of 30 to 50 years with past medical history of diabetes mellitus presented with abdominal pain. The patient was having a two week history of abdominal pain and dysuria. The patient became unresponsive at home and EMS brought the patient to the emergency room. At admission, the patient was hypoxic, tachycardic and hypotensive. Physical exam revealed diffusely tender abdomen and, coarse breath sounds. Laboratory values were significant for elevated lactate (11.8 mmol/L) and thrombocytopenia (112 × 10^3^ /ul) (Table S1). CT imaging of the chest and abdomen showed extensive pulmonary septic emboli, prostate abscesses, right pyelonephritis, multiple hepatic abscesses and thrombus within inferior vena cava (Figure 1). The patient was subsequently admitted to the intensive care unit and managed with appropriate fluid resuscitation, vasopressors, mechanical ventilator support, meropenem and heparin drip.

**Figure 1:**
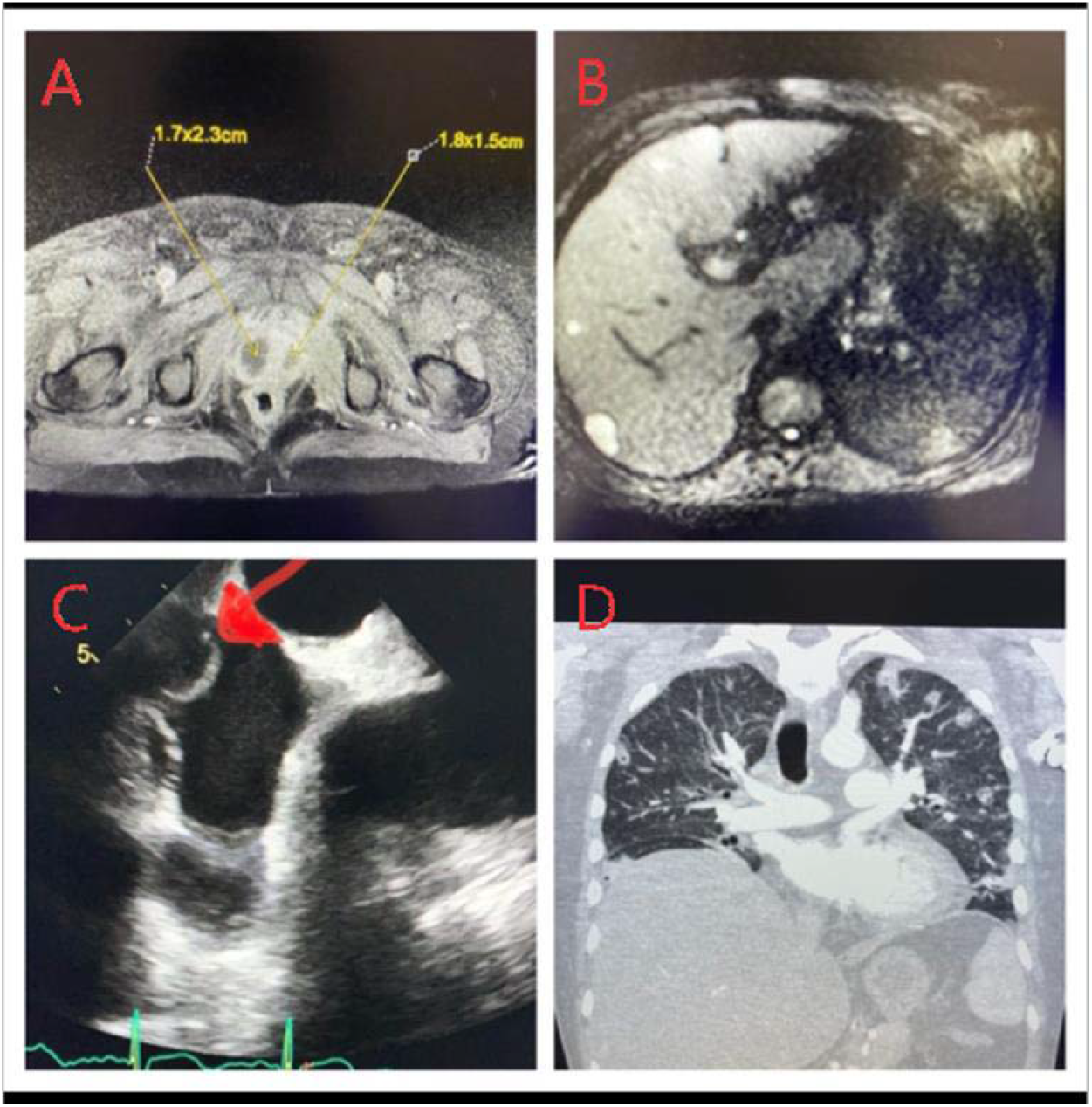
Metastatic septic infection by a hypervirulent Klebsiella pneumoniae (2020CK-00441). Panel A: MRI Pelvis showing bilateral prostatic abscesses measuring 1.7×2.3cm on right and 1.8×1.5cm on left. Panel B: MRI Abdomen series showing multiple small hepatic abscesses throughout the liver. Panel C: Trans-esophageal echocardiogram showing a clot traversing the right atrium from the inferior vena cava. Panel D: CT imaging of chest showing multiple septic emboli throughout bilateral lung fields.

Blood cultures grew *Klebsiella pneumoniae* (2020CK-00441). Transesophageal echocardiogram showed a clot in the inferior vena cava projecting into the right atrium and did not show vegetations or paravalvular abscess (Figure 1). Patient was extubated after three days and weaned off vasopressor support within one week. MRI of the abdomen and pelvis showed multiple hepatic abscesses and was negative for gall stones, psoas abscess or pelvic osteomyelitis (Figure-1). MRI of the brain, cervical, thoracic and lumbar spine were negative for abscesses and stroke and the patient was discharged after 16 days in stable condition.

#### b) Case #2

A patient in the age group of 30 to 50 years with diabetes mellitus and hypertension presented with right abdominal pain and back pain. The patient had a syncopal episode 10 days prior to presentation. The patient then started developing headache, poor appetite, dizziness and back pain necessitating the patient to come to the emergency room. On admission the patient was afebrile, tachycardic (103 /min), abdomen was soft, neck was supple, and lungs had good air entry without any rales. The patient laboratory values were significant for elevated neutrophil band cells (42%) and thrombocytopenia (50 × 10^3^ /ul)(Table S1). CT head without contrast did not show any acute abnormality. CT imaging of the chest and abdomen showed septic emboli in the lung, enlarged liver with high suspicion for a liver abscess and hepatic vein thrombosis. The patient blood pressure was 116/76 mm Hg, ABG showed normal pH, and lactate was mildly elevated (2.3 mmol/L). The patient was admitted to the internal medicine team and started on Piperacillin/Tazobactam, long-acting insulin and maintenance fluids. In view of severe thrombocytopenia, full dose anti-coagulation was not started.

Six hours from presentation, the patient became hypotensive and encephalopathic. Focal weakness or seizure like activity were not observed. Laboratory values were significant for worsening lactic acidosis (8.8 mmol/L) and worsening thrombocytopenia (30 × 10^3^ /ul). Patient care was escalated to the intensive care team. Appropriate fluid resuscitation, insulin drip, mechanical ventilation and vasopressors were started. Unfortunately, 20 hours from presentation the patient had cardiac arrest and could not be resuscitated even after more than an hour of cardiopulmonary resuscitation. Two days later, the blood cultures were positive for *Klebsiella pneumoniae* (2021CK-00720) while urine cultures were negative.

### B. Clinical Laboratory

Antibiotics sensitivity testing was performed on both of the isolates as described in the methods. Multi-drug resistance or carbapenam resistance was not observed in both the isolates. The detailed antibiotic sensitivities and minimal inhibitory concentration profiles using routine laboratory testing and alternative Sensititre GNX2F panel testing for both isolates are provided in Supplementary Table 2 and 3. Additionally, carbapenemases were not detected by modified carbapenem inactivation method (mCIM) testing.

### C. Basic Science

#### a) Whole Genome Sequencing

Hybrid assembly of 2020CK-00441 resulted in one closed chromosome sequence, two closed plasmid sequences, and no known incomplete plasmid sequences (Figure 2). PGAP identified 5,338 genes and 130 pseudogenes in the assembly. Details of the annotation are provided in Supplementary data. Loci and genes associated with hypervirulence were identified on the chromosomal sequence resembling the ICEKp10 mobile element and p2020CK-00441_1.

**Figure 2:**
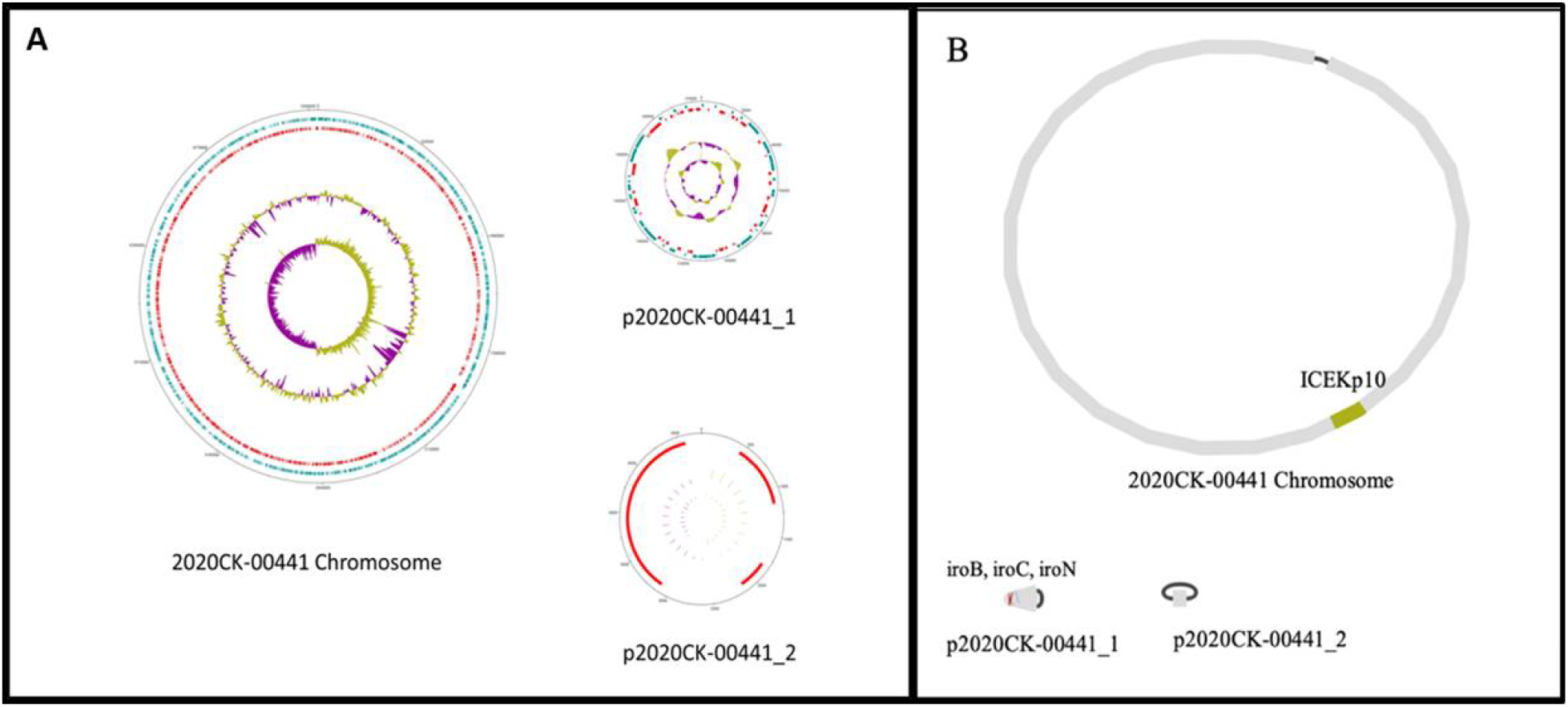
A) Complete genome sequence of 2020CK-00441 with circular chromosome and 2 plasmids with expected gene distribution and GC skew as show by Artemis. B) Bandage graph of assembly with selected virulence factor locations colored.

Although long-read sequencing was not performed on 2021CK-00720, both of the strains were given a score of five for virulence by Kleborate analysis which shows high probability of the isolates to be hvKp strains (Table 1 and Supplemental material 2). Both of the isolates did not have any genomic evidence of multi-drug resistance or carbapenam resistance (Supplementary Data). 2020CK-00441 had ybt1-clb2 profile while 2021CK-00720 had ybt17-clb3 profile. Both the isolates had virulence genes encoding aerobactin (iuc1), salmochelin (iro1) and rmpADC loci which are frequently present in pK2044 like plasmids in hvKp strains. Presence of rmpADC loci in both the isolates suggests that both are likely to have a hypermucoviscous phenotype.

**Table 1:** Summary of Kleborate Analysis:

| Isolate | 2020CK-00441 | 2021CK-00720 |
| --- | --- | --- |
| Species | Klebsiella pneumoniae | Klebsiella pneumoniae |
| ST | ST23 | ST65 |
| Virulence Score | 5 | 5 |
| Resistance Score | 0 | 0 |
| Number of Drug Resistance Genes | 0 | 0 |
| Yersiniabactin | ybt 1; ICEKp10 | ybt 17; ICEKp10 |
| Colibactin | clb 2 | clb 3 |
| Aerobactin | iuc 1 | iuc 1 |
| Salmocheilin | iro 1 | iro 1 |
| rmpADC | rmp 1; KpVP-1 | rmp 1; KpVP-1 |
| rmpA2 | rmpA2_6-60% | rmpA2_5-54% |
| wzi | wzi1 | wzi72 |
| K_locus | KL1 | KL2 |
| O_type | O1 | O1 |

Legend: Summary of kleborate analysis showing that both the isolates are highly likely to be hvKp (Virulence score = 5) and unlikely to be multi-drug resistant (Resistance Score = 0)

#### b) Comparison of p2020CK-00441_1 and pK2044

p2020CK-00441_1 is a non-self-transmissible IncFIBK type virulence plasmid. When compared to previously described pK2044 plasmid, it is nearly identical (99.98% identity and 98% coverage) (Figure 3). Due to the potential clinical significance of the role of the plasmid in HvKp phenotype we performed further analysis and compared the plasmid sequence with historic sequence pK2044 sequence as well as the top ten BLAST hits (Figure 3). Although we see vast amounts of synteny of p2020CK-00441_1 and other closely related plasmids, the gene architecture is unique and undescribed previously. The significance of this unique plasmid on the ability to be hypervirulent is unknown and needs further research.

**Figure 3:**
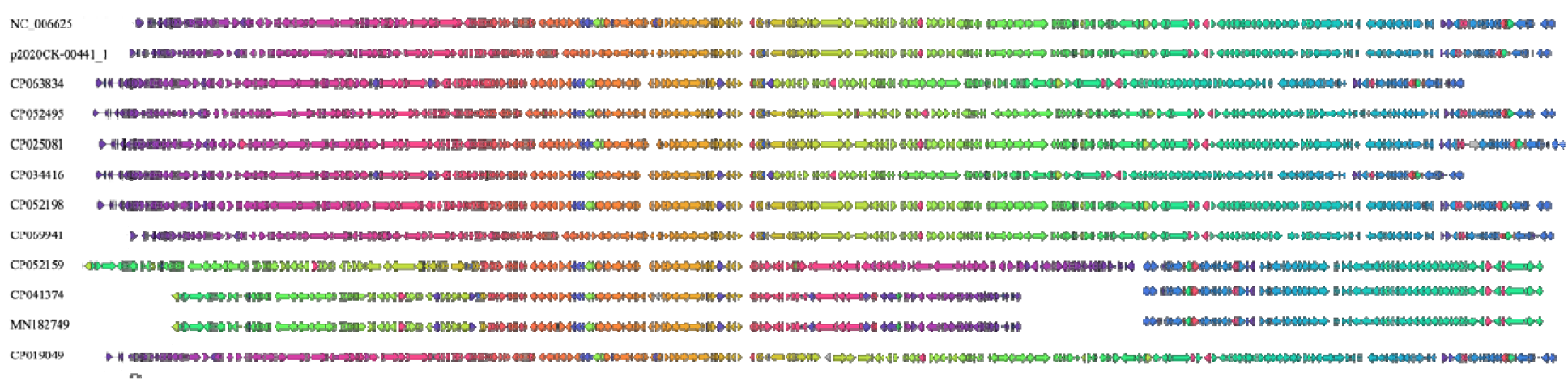
Synteny alignment of p2020CK-00441_1, NC_006625/ pK2044 and the top 10 similar plasmids identified via blast. Synteny groups share colors. Plasmid sequences were wrapped around manually for clarity.

The second smaller plasmid, p2020CK-00441_2 is nearly identical to prior described ColRNAI type *Klebsiella* plasmids, pUUH239.1 and pBK13048_7 (99% identity and 100% coverage). These plasmids encode for plasmid mobilization proteins and phage resistance proteins and have no known role in HvKp phenotype.

#### c) ICE regions of 2020CK-00441

2020CK-00441 contains the ICEKp10 mobile element in its chromosome, which is typical of ST23 type Klebsiella [29, 30]. When compared with KY454634, the arrangement of genes is almost an exact match, with the exception of a hypothetical protein with similarity to IS5 family transposase IS903 (Figure 4). It is unclear if this gene insertion impacts hypervirulence. 2021CK-00720 is also expected to contain the ICEKp10 mobile element by kleborate analysis, but this was not able to be confirmed due to the limitations of short-read Illumina sequencing.

**Figure 4:**
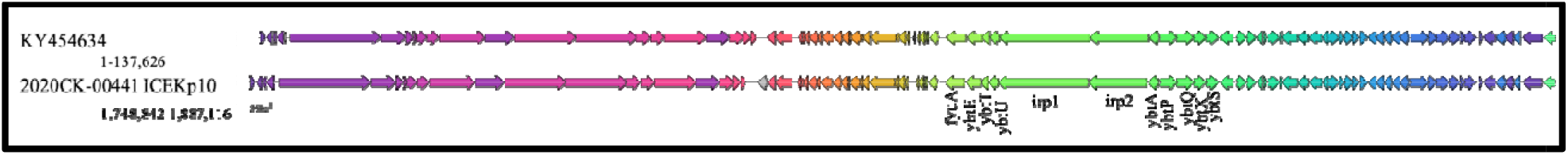
Synteny alignment of the ICEKp10 region of 2020CK-00441 with annotated hypervirulent genes. Synteny groups share colors.

Homology searches revealed an additional potential ICE element in 2020CK-0441 which is similar to *E*.*coli* ICEEc2. This region encodes for hypothetical genes and Type IV secretion system. None of these genes have been implicated in contributing to hvKp phenotype thus far.

#### d) Comparison of 2020CK-00441 and 2021CK-00720

Although no known hvKp cases were observed previously in the past decade at HCA Houston Healthcare, both 2020CK-00441 and 2021CK-00720 presented a few weeks a part. In order to determine if these two cases were due to related exposures or independent occurrences, the core genomes of 2020CK-00441 and 2021CK-00720 were compared. 2020CK-00441 and 2021CK-00720 shared 4,577 out of 5,646 genes identified, but these shared genes differed by 29,575 SNPs. Additionally, 2021CK-00720 was predicted to be ST65 (Table 1). This suggested that a) the isolates were not closely related, b) unlikely to be part of same outbreak and c) there may be more than one strain of hvKp present in the community around our hospital.

#### e) Phylogenetic tree analysis

In order to determine if the two observed cases were similar to other hvKp studies reported from other continents, a phylogenetic tree was created with the ST types of both of our isolates with the prior reported hvKp Bioprojects (Figure 5). Surprisingly, the genomic sequences of both of the strains were not closely related to any of the other hvKp sequence submitted to NCBI thus far. This suggests that the two US isolates are distinct from the isolates in other continents and have the potential to have varying clinical and microbiological profiles.

**Figure 5:**
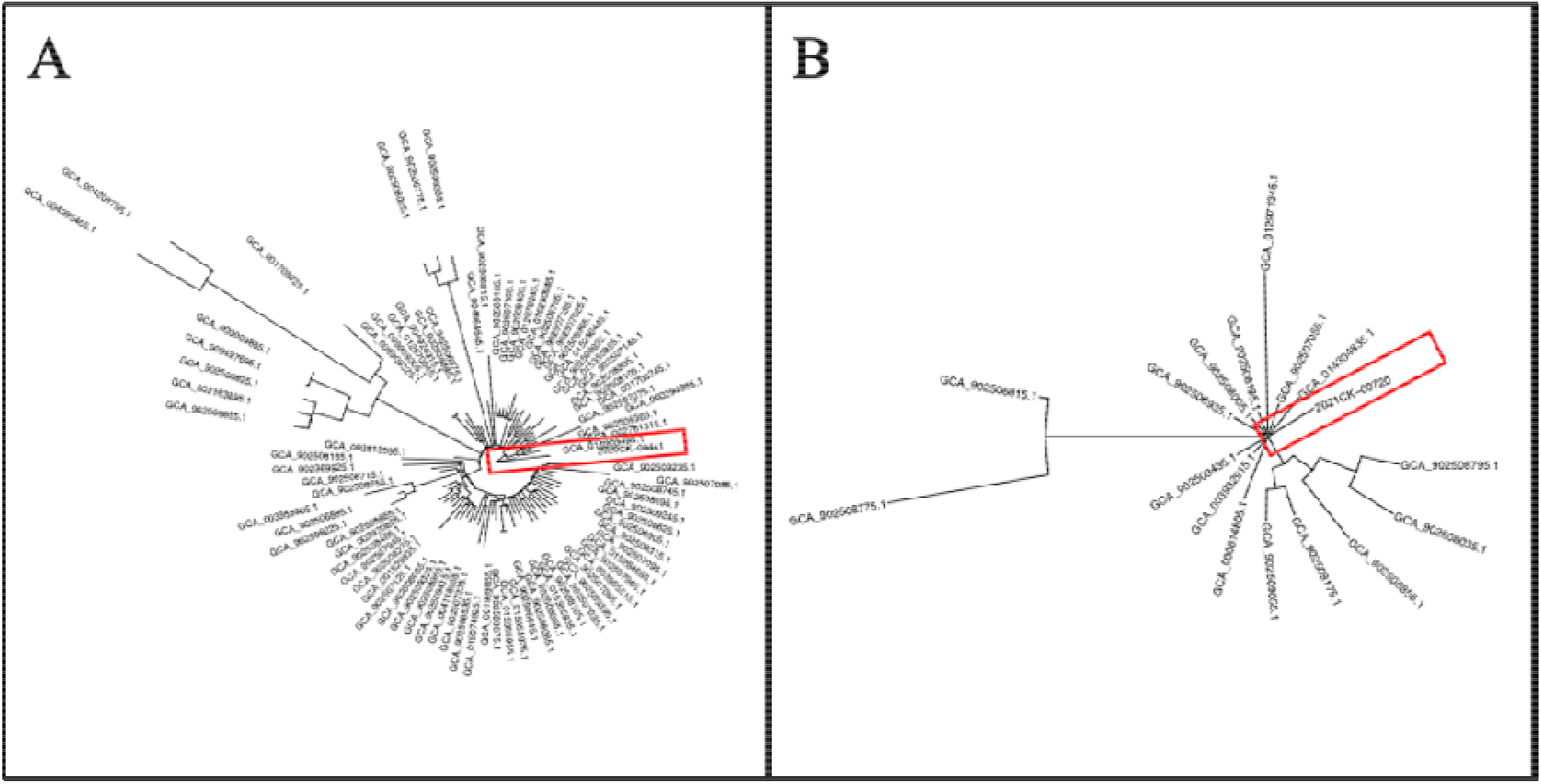
Phylogenetic trees of A) 2020CK-00441 (highlighted with red box) with available ST23 genomes and B) 2021CK-00720 (highlighted with red box) with available ST65 genomes.

### D. Epidemiology

In addition to review of hospital chart for social history, phone survey of patient and/or patient family to identify occupational or travel related exposures was performed before the submission of this report. One of the patients did not have any known sick contacts, occupational exposure and did not have any international travel while the other patient had significant remote history of international travel.

## DISCUSSION

In this report we were able to show that the two infections were caused by vastly different hvKp bacterial strains in two patients from significantly different demographic backgrounds. Definitive conclusions regarding optimal clinical management cannot be made with such a small sample size. A detailed review of the two cases was performed by the physicians of our team. The general principles that we would recommend to clinicians in managing these patients include, a) high clinical suspicion of these infections b) appropriate admission to critical care unit c) initial broad spectrum antibiotics d) aggressive imaging to identify foci of infection and e) therapeutic anti-coagulation.

High clinical suspicion to identify these infections is vital. Physicians in United States would need to include hvKp metastatic septic infections in the differential diagnosis of all patients with septic emboli especially in young poorly controlled diabetics. Patients with encephalopathy, thrombocytopenia and high number of immature neutrophils (band cells) may have poor prognosis and may require intensive care admission. CT-imaging of chest, abdomen and pelvis may help to identify large abscesses and extent of the disease. MRI of the abdomen and pelvis may be needed to delineate the abscesses in the abdomen and also identify pelvic bone osteomyelitis. Dilated eye exam, lumbar puncture and MRI of the brain and spine may be required for identifying neurological septic infections. Both the patients showed thrombophlebitis and clots in the major veins of the abdomen which likely led to septic emboli in the lung. Therapeutic anti-coagulation whenever feasible may be beneficial.

The two patients described in this report do not share any common epidemiological risk factors or links. With no known exposure or travel history, one of the patients appears to have aquired the disease in the community. The other patient’s infection cannot be exclusively attributed to community spread as the patient had significant international travel history.

Chou et al are the only publication describing hvKp in Texas that we could identify from literature search, screened 64 Klebsiella pneumoniae isolates for putative hvKp genes rmpA and magA in two major hospitals of Houston, Texas (about 25 miles south of our hospital) [18]. Only one of the four isolates was associated with liver abscesses while the others did not cause clinically relevant infections. The cases did not have a metastatic septic infection and the isolates did not undergo WGS. Whether these isolates indicate the presence of strains causing milder disease which have now evolved to cause much more devastating disease or whether the isolates we saw in our patients were new and introduced recently into the community is unknown at this time.

In this report we show that both 2020CK-0041 and 2021CK-00720 have genomic content necessary to make additional siderophores like yersiniabactin, aerobactin and salmochelin. Highly virulent strains in other bacterial species have been shown to have higher expression of siderophore producing genes when compared to less virulent strains of the same species [31, 32]. All of these factors are postulated to give these strains an advantage to obtain iron and counteract the innate defense mechanisms of the human body. Whether these siderophores are actually synthesized and if these siderophores play a role in the causation of the metastatic septic disease would need future studies.

From a public health perspective, it is alarming that two unrelated hvKp infections were discovered in a few weeks from each other in a single hospital. This suggests that there may be more hidden hvKp infections that are currently going undetected in the U.S. healthcare system. Surveillance of these organisms and hypervirulence factors with WGS is needed to adequately inform response and infection prevention. The recent addition of *Klebsiella* hypervirulence genetic markers on the list of public health alerts reportable by laboratories within the ARLN, is a timely and welcome inclusion.

The location of our hospital as one of the major hospitals serving the communities who live close to and work in the Houston Intercontinental Airport makes it possible that we may be seeing the index cases of locally adapted hvKp strains capable of causing devastating metastatic septic infections. By transfer of relatively short genomic sequences there is a potential for emergence of a large number of locally adapted hvKp strains. To understand the rate of emergence and spread of problematic strains, a geography based detailed catalog of the genomic sequences of both the cKP and hvKp is essential. A seamless “patient to laboratory to bench to community” translational approach with close collaboration of physicians, clinical microbiologists, basic science scientists, epidemiologists and health department is needed to tackle this emerging problem.

## Data Availability

All of the genome sequences have been submitted to the STA database

