## Supplemental data for "Emergence of Severe Metastatic Hypervirulent *Klebsiella pneumoniae* infections in Texas"

Supplemental Methods:

Blood cultures

Initially blood cultures were collected in Bactec lytic bottles, which contains soybean-casein digest broth, yeast, amino acids, sugar, vitamins and sodium polyanetholsulfonate along with a detergent that lyses red and white blood cells in an attempt to increase yield. These are then placed in the Bactec FX system; the system works through identifying CO2 released by microorganisms. The CO2 release causes a dye trigger on the bottle which is then detected by fluorescence which the Bactec machine flags as positive. Once the blood culture returns positive they are Gram-stained and cultured to a blood, MacConkey agar and chocolate agar plates for growth. Once growth from the sub-culture is identified, the colonies are then emulsified, and a Microscan Walkaway is used for automated identification and sensitivities.

Identification and antibiotic sensitivity testing

(i) Microscan Walkaway system

Miroscan is able to identify the organism through use of fluorogenic panels. First the colony is diluted with medium then poured into a loading tray for the panels. This inoculum is then pipetted into individual wells with antimicrobials on the plate designed for Gram negative pathogens specifically to give Minimal Inhibitory Concentration (MIC). For our patient samples, a Combo 67 plate was used. After incubation, the growth is determined by the amount of light passing through this well then placed into an algorithm to give MIC breakpoints and likely pathogen.

(ii) MALDI-TOF and Sensititre GNX2F panels (Thermo-Fisher Scientific)

Bacterial isolates are identified to genus and species level using matrix-assisted laser desorption/ionization (MALDI-TOF), a rapid identification system. The Sensititre system is a micro broth dilution method in dried plate format for determining antibiotic susceptibilities. GNX2F panels are inoculated with a standardized suspension of the bacteria. Panels are incubated at 34-36 C for 18-24 hours in a non-CO2 incubator. After incubation, MIC results are read using the Sensititre manual viewer

Modified Carbapenem Inactivation Method (mCIM):

The Modified Carbapenem Inactivation Method (mCIM) is a phenotypic method for detection of carbapenemase production by bacteria. A standardized suspension of the bacteria is incubated in broth for a minimum of 4 hours with a 10 ug meropenem-impregnated disk. Potential inactivation is then analyzed by placing the carbapenem disk on a lawn of a carbapenem-susceptible indicator stain of *Escherichia coli*, typically ATCC 25922 and incubating overnight. No zone or a small zone of inhibition around the disk, indicates the test isolate is a carbapenemase-producer.

Supplemental Table 1:

Labs at presentation :

| **Labs** | **Case 1** | **Case 2** |
| --- | --- | --- |
| **White Blood Cells** | *26.2 x 10^3^/ul* | *7.4 x 10^3^/ul* |
| **Hemoglobin** | *8.8 g/dl* | *14.4 g/dl* |
| **Platelets** | *112 x 10^3^/ul* | *52 x 10^3^/ul* |
| **Sodium (uncorrected)** | *129 mmol/L* | *126 mmol/L* |
| **Chloride** | *97 mmol/L* | *91 mmol/L* |
| **Potassium** | *3.3 mmol/L* | *3.4 mmol/L* |
| **Ionized Calcium** | *1.05 mmol/L* | *1.09 mmol/L* |
| **Magnesium** | *1.8 mg/dl* | *1.8 mg/dl* |
| **Phosphorous** | *3.4 mg/dl* | *2.4 mg/dl* |
| **Blood total Carbon dioxide** | *12 mmol/L* | *22 mmol/L* |
| **Glucose** | *271 mg/dl* | *501 mg/dl* |
| **Blood Urea Nitrogen** | *24 mg/dl* | *42 mg/dl* |
| **Creatinine** | *1.0 mg/dl* | *1.3 mg/dl* |
| **INR** | 2.7 | 1.2 |
| **Thromboplastin Time Partial** | 31.2 seconds | 34.6 seconds |
| **Aspartate Transaminase (AST)** | 191 U/L | 70 U/L |
| **Alanine Aminotransferase (ALT)** | 63 U/L | 132 U/L |
| **Alkaline Phosphatase** | 751 U/L | 182 U/L |
| **Total bilirubin** | 3.3 mg/dl | 2.0 mg/dl |
| **Bilirubin Conjugated** | 1.2 mg/dl | 0.1 mg/dl |
| **Total Protein** | 5.4 g/dl | 6.8 g/dl |
| **Albumin** | 2.2 g/dl | 3.6 g/dl |
| **Lactic Acid** | 11.8 mmol/L | 2.3 mmol/L |
| **D-Dimer** | 7225 ng/ml | Not Available |
| **Point of Care ABG, pH** | 7.42 | 7.41 |
| **Point of Care ABG, pCO2** | 18.8 mmHg | 32.4 mmHg |
| **Point of Care ABG, pO2** | 81.1 mmHg | 75.8 mmHg |
| **Troponin-I** | <.012 ng/ml | 0.018 ng/ml |
| **Hemoglobin A1C** | 10.4 % | 13.0 % |
| **Ammonia** | 32 | <9 umol/L |
| **CK** | <30 | <30 U/L |
| **COVID Antigen** | Negative | Negative |
| **HIV 1&2 Antibody** | Negative | Not Available |
| **Hepatitis A, B, C antibodies** | Negative | Negative |

Supplemental Table 2:

Antibiotic Sensitivity for the two isolates:

|  | Case – 1 Sensitivity | Case 1 MIC (mg/L) | Case – 2 Sensitivity | Case 2 MIC (mg/L) |
| --- | --- | --- | --- | --- |
| Gentamicin | Sensitive | <=2 | Sensitive | <=2 |
| Ampicillin | Resistant | >16 | Resistant | >16 |
| Cefazolin | Sensitive | <=4 | Sensitive | <=4 |
| Tobramycin | Sensitive | <=2 | Sensitive | <=2 |
| Tetracycline | Sensitive | <=2 | Sensitive | <=2 |
| Trim/ Sulfa | Sensitive | <=2/38 | Sensitive | <=2/38 |
| Amikacin | Sensitive | <=8 | Sensitive | <=8 |
| Ceftriaxone | Sensitive | <=1 | Sensitive | <=1 |
| Ertapenam | Sensitive | <=0.5 | Sensitive | <=0.5 |
| Cefuroxime | Sensitive | <=4 | Intermediate | 16 |
| Cefepime | Sensitive | <=1 | Sensitive | <=1 |
| Aztreonam | Sensitive | <=4 | Sensitive | <=4 |
| Amp/Sulbactam | Sensitive | 8/4 | Sensitive | 8/4 |
| Piperacil/Tazo | Sensitive | <=16 | Sensitive | <=16 |
| Meropenem | Sensitive | <=1 | Sensitive | <=1 |

Interpretations based on CLSI M100-S24*

*CLSI M100-S24: Performance Standards for Antimicrobial Susceptibility Testing; Twenty-Fourth Informational Supplement. CLSI document M100-S24. Wayne, PA: Clinical and Laboratory Standards Institute; 2014

Supplementary Table 3

|  | Case – 1 Sensitivity | Case 1 MIC (mg/L) | Case – 2 Sensitivity | Case 2 MIC (mg/L) |
| --- | --- | --- | --- | --- |
| Amikacin | Susceptible | <=4 | Susceptible | <=4 |
| Aztreonam | Susceptible | <=2 | Susceptible | <=2 |
| Piperacil/Tazo | Susceptible | <=8/4 | Susceptible | <=8/4 |
| Trim/ Sulfa | Susceptible | <=0.5/9.5 | Susceptible | <=0.5/9.5 |
| Gentamicin | Susceptible | <=1 | Susceptible | <=1 |
| Cefepime | Susceptible | <=2 | Susceptible | <=2 |
| Tobramycin | Susceptible | <= 1 | Susceptible | <= 1 |
| Meropenem | Susceptible | <=1 | Susceptible | <=1 |
| Cefotaxime | Susceptible | <= 1 | Susceptible | <= 1 |
| Tigecycline | Susceptible | <=0.25 | Susceptible | <=0.25 |
| Ertapenem | Susceptible | <=0.25 | Susceptible | <=0.25 |
| Imipenam | Susceptible | <=1 | Susceptible | <=1 |
| Doripenam | Susceptible | <=0.12 | Susceptible | <=0.12 |
| Colistin | Intermediate | <=0.25 | Intermediate | <=0.25 |
| Ceftazidime | Susceptible | <=1 | Susceptible | <=1 |
| Minocycline | Susceptible | <=2 | Susceptible | <=2 |
| Doxycycline | Susceptible | <=2 | Susceptible | <=2 |

Supplemental Table 4: Genomic Features of 2020CK-00441:

|  | 2020CK-00441 |
| --- | --- |
| Genes | 5338 |
| CDS (total) | 5216 |
| Genes (coding) | 5086 |
| Genes (RNA) | 122 |
| rRNA (5S, 16S, 23S) | 9, 8, 8 |
| tRNA | 86 |
| ncRNA | 11 |
| Pseudo Genes | 130 |
| Plasmids | 2 |

Of the 130 total pseudogenes there were 63 that were due to incomplete predicted gene regions, 39 due to a predicted internal stop and 64 due to frameshifts (32 of the 130 pseudogenes had multiple problems leading to more pseudogene causing events than total number of pseudogenes). 2021CK-00720*: The sequence of 2021CK-00720 is at a contig stage and not complete hence not included in this table.
